# Chain of Survival Complexities and Barriers in the Muslim Community

**DOI:** 10.64898/2026.03.05.26347762

**Authors:** Heather Liffert, Maha Shoaib, Shestruma Parajuli, Brian Meier, Leslie Chavez, John C. Perkins

**Affiliations:** Compress and Shock Foundation, Roanoke, VA 24018; Edward Via College of Osteopathic Medicine, Blacksburg, VA 24060; Virginia Tech Carilion School of Medicine, Roanoke, VA, 24016; Department of Emergency Medicine, Carilion Clinic, Roanoke, VA 24014

## Abstract

**Background:** Out-of-hospital cardiac arrest (OHCA) survival depends on timely bystander cardiopulmonary resuscitation (CPR) and quick defibrillation via automated external defibrillator (AED). However, access to CPR education and willingness to intervene are not equitably distributed. Within the Muslim community, intersecting religious identity, language, immigration-related concerns, and other social determinants of health may affect CPR/AED education, bystander response, and ultimately OHCA outcomes, underscoring the need for culturally responsive, faith-based training models.

**Methods:** A survey based cross sectional study was conducted to evaluate the perceived barriers to emergency response and lay rescuer cardiopulmonary resuscitation (CPR). Individuals aged 13 years and older were recruited between January and June 2025 through convenience sampling at free, non-certification public CPR/AED classes, where participants self-reported demographic characteristics and barriers to calling 9-1-1 or initiating CPR. Analyses compared Muslim and non-Muslim participants using Fisher exact tests and multivariable logistic regression models adjusted for demographic and socioeconomic factors, with results reported as odds ratios (OR) and 95% confidence intervals (CI).

**Results:** Of the 651 surveys collected, 33% of participants identified as Muslim, and 46% reported no prior CPR/AED training, with a higher proportion among Muslim respondents (57% vs 41%). Religion was significantly associated with some perceived barriers, with Muslim participants more likely to report law enforcement as a barrier to calling 9-1-1 (OR: 0.53 for non-Muslims vs Muslims, p=0.04) and less likely to report “no problem” starting CPR (OR: 0.91, p=0.04). Race and gender also influenced barriers, with non-white and female participants more likely to report immigration status, language, cost, and concern for violence as barriers to initiating CPR or calling 9-1-1.

**Conclusion:** Muslim participants were more confident in performing CPR, but reported less confidence in calling 9-1-1, revealing gaps in emergency response readiness. This emphasizes the importance of culturally adapted CPR/AED training that addresses specific barriers within faith-based communities and to strengthen all links of the chain of survival.

## INTRODUCTION

Out-of-hospital cardiac arrest (OHCA) remains a leading cause of mortality globally. A central concept in responding to OHCA is the “chain of survival”. The chain of survival is a sequence of time-sensitive events or “links”: early recognition of cardiac arrest and activation of the emergency-response system; immediate, high-quality cardiopulmonary resuscitation (CPR); rapid defibrillation; basic and advanced EMS; and advanced life support and post arrest care.^1^ Bystander delivery of CPR occurs in less than 40% of OHCA victims.^2^ This may be due to a misunderstanding or lack of knowledge of the capabilities of CPR, resuscitative measures, and other healthcare interventions.^3^ Health inequities and disparities further affect the rate of survival, with disproportionately lower rates of bystander CPR seen in areas of socioeconomic deprivation and ethnic minority groups.^4^

The 2025 AHA CPR guidelines further report that survival and neurological outcomes after cardiac arrest are significantly worse in minoritized racial and ethnic groups compared with non-Hispanic white individuals.^5^ In addition, racial and ethnic differences report non-white individuals being less likely to receive bystander CPR.^6–8^ A 2022 study demonstrated that regardless of location (at home or in public) individuals who were Black or Hispanic were 10-20% less likely to receive bystander CPR compared to their White counterparts.^6^ On a global scale, women are less likely to receive bystander CPR compared to men.^9^ Research on language barriers highlights underutilization of 9-1-1, delays in care, and survival outcomes for non-English speakers.^10–12^

These disparities are in part due to social determinants of health (SDOH) or nonmedical factors that influence health outcomes.^13^ Lack of equal access to CPR education is affected by each of these. In studies that have examined CPR education, there are lower rates of accessing education among individuals who are women, non-white, older, and of a lower socioeconomic status.^14–16^ Several factors have been identified that may represent barriers to CPR education/access, as well as activation of the chain of survival. Examples include language barrier, class cost, fear/distrust of law enforcement, fear over immigration status, liability concerns, and fear of causing harm to the victim.^17–20^ These barriers, shaped by SDOH, provide an opportunity to tailor CPR/AED education in specific communities.

Models of CPR/AED education that are developed to provide education CPR training in faith institutions increase accessibility to marginalized communities who are often excluded from this education.^4^ However, few studies have explored how religious identity and systemic barriers shape CPR/AED education and bystander action. The Muslim community represents one such population disproportionately affected. It is important to distinguish between Islam, the religion itself (including its teachings, practices, and core beliefs) and Muslims, the diverse population of individuals who practice the faith. The term Islamic is used as an adjective to describe a characteristic of Islam.^21^ Barriers to equitable emergency medical services (EMS), stemming from the complexity of their intersectional identities, are one such disparity faced by this community.^22^ “Mosque lifesaver training” and other educational models have provided CPR training directly in mosques, utilized multilingual translation, or provided trainings for free to limit barriers.^23–24^

Significant gaps remain in understanding how intersectional identities, such as religion, influence access to CPR/AED delivery and education. This study aims to examine intersecting factors in order to inform the development of a culturally responsive CPR/AED education model that addresses the disparities in OHCA outcomes faced by Muslim communities. We hypothesize that multiple barriers may exist in the Muslim learner population that requires tailored education in order to reduce hesitancy.

## METHODS

This study consisted of cross-sectional, survey-based research. A survey instrument (Appendix A) was designed to assess the knowledge, comfort, and perceived barriers to activating the 9-1-1 system and performing lay rescuer CPR. IRB approval and exemption was determined through Carilion Clinic IRB.

### Study Sample

Participants were recruited in person between January 2025 - June 2025 using a convenience sampling strategy at free, non-certification, public CPR/AED classes. All persons aged 13 and older were eligible to take the survey. Surveys were distributed as participants arrived prior to the class start and were optional. Surveys did not collect private health information (PHI) and were completed on paper and stored using Research Electronic Data Capture (REDCap) at Virginia Tech Carilion School of Medicine.^25–26^ The data that support the findings of this study are available on request from the corresponding author upon reasonable request.

### Survey Design and Measures

Demographic information (zip code, age range, gender, race/ethnicity, education level, income) was collected via the survey instrument and was self-reported. Survey items related to perceived barriers to calling 9-1-1 or starting CPR asked participants to consider the following barriers: law enforcement, immigration status, cost, fear of doing something wrong, language barrier, concern for violence. In addition, participants had the option to mark “other” or no barrier to calling 9-1-1/starting CPR.

### Statistical Analysis

All analyses and data management were conducted using R 4.3.1.^27^ Survey responses were directly compared between religions (Muslim vs. Non-Muslim) using Fisher tests. To analyze the association between religions and perceived barriers, a model was built for each of the barriers. Models included adjustments for variables that had statistically significant Fisher test results. Each model was adjusted for primary language, gender, age group, relationship status, race, education level and household income before being run against the outcome of religion. For all models, the reference values were based upon previous literature to compare any relevant results to those of past studies.

Models were built using the “glm” function from the package “stats”.^28^ Regression coefficient estimates were exponentiated, and the 95% confidence intervals (CI) were calculated. Results of the regressions are presented as odds ratios (OR) with 95% CI and p-values.

## RESULTS

### Survey Response

As survey participation was voluntary, response rates varied across events. Upon entering an event, participants were asked if they were willing to take a survey. If they agreed, they were handed a survey and a writing utensil with instructions to return surveys when complete. Surveys were collected at 17 different events. The average number of learners at events was approximately 77 people and the average survey response rate was approximately 68.8%.

#### Descriptive Statistics

A total of 651 surveys were collected. Of these, 214 participants indicated Muslim as their religion (33%) and 437 indicated other religions (67%). Table 1 presents demographic information of survey participants. Overall, females made up a higher percentage of survey participants (61%) compared to males (38%). 46% of participants indicated they had not received prior CPR/AED training - 57% of these were Muslim and 41% were non-Muslim. Fisher test results showed that all barriers to starting CPR, except for concern for violence, had a significant association (p < 0.05) with religion (Table 2). Barriers to calling 9-1-1 that were significant as a result of the Fisher test between religion and barriers were cost and law enforcement (Table 2).

**Table 1.** Sociodemographic Characteristics of Survey Participants.

| Table 1. Sociodemographic Characteristics of Survey Participants |  |  |  |  |
| --- | --- | --- | --- | --- |
| Characteristic | Overall<br>(N = 651) | Muslim<br>(N = 214) | Non-Muslim<br>(N = 437) | p-value |
| Primary Language |  |  |  | **<0.001** |
| English | 545 (84%) | 184 (86%) | 361 (83%) |  |
| Spanish | 61 (9.4%) | 0 (0%) | 61 (14%) |  |
| Multilingual | 31 (4.8%) | 18 (8.4%) | 13 (3.0%) |  |
| Other | 11 (1.7%) | 11 (5.1%) | 0 (0%) |  |
| I choose not to answer | 3 (0.5%) | 1 (0.5%) | 2 (0.5%) |  |
| Gender |  |  |  | **<0.001** |
| Man | 250 (38%) | 115 (54%) | 135 (31%) |  |
| Woman | 396 (61%) | 98 (46%) | 298 (68%) |  |
| Non-binary/gender non-conforming | 2 (0.3%) | 0 (0%) | 2 (0.5%) |  |
| I choose not to answer | 2 (0.3%) | 1 (0.5%) | 1 (0.2%) |  |
| Age |  |  |  | **<0.001** |
| 13-25 years old | 156 (24%) | 89 (42%) | 67 (15%) |  |
| 26-40 years old | 106 (16%) | 46 (21%) | 60 (14%) |  |
| 41-55 years old | 146 (22%) | 53 (25%) | 93 (21%) |  |
| 56-70 years old | 183 (28%) | 19 (8.9%) | 164 (38%) |  |
| 71-85 years old | 52 (8.0%) | 6 (2.8%) | 46 (11%) |  |
| over 85 years old | 4 (0.6%) | 0 (0%) | 4 (0.9%) |  |
| I choose not to answer | 4 (0.6%) | 1 (0.5%) | 3 (0.7%) |  |
| Relationship Status |  |  |  | **<0.001** |
| Single | 245 (38%) | 97 (45%) | 148 (34%) |  |
| Married | 322 (49%) | 106 (50%) | 216 (49%) |  |
| Living with partner | 15 (2.3%) | 1 (0.5%) | 14 (3.2%) |  |
| Separated | 7 (1.1%) | 0 (0%) | 7 (1.6%) |  |
| Divorced | 23 (3.5%) | 1 (0.5%) | 22 (5.0%) |  |
| Widowed | 28 (4.3%) | 3 (1.4%) | 25 (5.7%) |  |
| Other | 2 (0.3%) | 1 (0.5%) | 1 (0.2%) |  |
| I choose not to answer | 9 (1.4%) | 5 (2.3%) | 4 (0.9%) |  |
| Race |  |  |  | **<0.001** |
| Asian or Pacific Islander | 4 (0.6%) | 1 (0.5%) | 3 (0.7%) |  |
| Black or African American | 121 (19%) | 104 (49%) | 17 (3.9%) |  |
| Hispanic or Latino | 252 (39%) | 33 (15%) | 219 (50%) |  |
| Native American or Alaskan Native | 91 (14%) | 4 (1.9%) | 87 (20%) |  |
| White or Caucasian | 121 (19%) | 32 (15%) | 89 (20%) |  |
| Multiracial or Biracial | 14 (2.2%) | 3 (1.4%) | 11 (2.5%) |  |
| Other | 31 (4.8%) | 28 (13%) | 3 (0.7%) |  |
| I choose not to answer | 16 (2.5%) | 9 (4.2%) | 7 (1.6%) |  |
| Highest Level of Education |  |  |  | **<0.001** |
| Less than high school | 76 (12%) | 40 (19%) | 36 (8.2%) |  |
| High school graduate/GED | 114 (18%) | 15 (7.0%) | 99 (23%) |  |
| Some college (no degree) | 89 (14%) | 18 (8.4%) | 71 (16%) |  |
| Vocational training | 21 (3.2%) | 1 (0.5%) | 20 (4.6%) |  |
| Associate's/bachelor's | 191 (29%) | 55 (26%) | 136 (31%) |  |
| Master's | 108 (17%) | 51 (24%) | 57 (13%) |  |
| PhD/MD/equivalent | 40 (6.1%) | 31 (14%) | 9 (2.1%) |  |
| I choose not to answer | 12 (1.8%) | 3 (1.4%) | 9 (2.1%) |  |
| Household Income |  |  |  | **0.007** |
| \$10,000 to less than \$20,000 | 15 (2.3%) | 4 (1.9%) | 11 (2.6%) | |
| Under \$5,000 | 23 (3.6%) | 6 (2.8%) | 17 (3.9%) | |
| \$20,000 to less than \$40,000 | 64 (9.9%) | 17 (8.0%) | 47 (11%) | |
| \$40,000 to less than \$60,000 | 67 (10%) | 13 (6.1%) | 54 (13%) | |
| \$5,000 to less than \$10,000 | 14 (2.2%) | 6 (2.8%) | 8 (1.9%) | |
| \$60,000 to less than \$80,000 | 62 (9.6%) | 16 (7.5%) | 46 (11%) | |
| \$80,000 or more | 181 (28%) | 74 (35%) | 107 (25%) | |
| I don't know | 57 (8.9%) | 28 (13%) | 29 (6.7%) |  |
| I choose not to answer | 161 (25%) | 49 (23%) | 112 (26%) |  |
| Religion |  |  |  | **<0.001** |
| Agnostic | 13 (2.0%) | 0 (0%) | 13 (3.0%) |  |
| Atheist | 1 (0.2%) | 0 (0%) | 1 (0.2%) |  |
| Christian | 332 (51%) | 0 (0%) | 332 (76%) |  |
| Jewish | 2 (0.3%) | 0 (0%) | 2 (0.5%) |  |
| Muslim | 214 (33%) | 214 (100%) | 0 (0%) |  |
| Other | 40 (6.1%) | 0 (0%) | 40 (9.2%) |  |
| I choose not to answer | 49 (7.5%) | 0 (0%) | 49 (11%) |  |
| Primary Care |  |  |  | **0.2** |
| Doctor's office (in-person visit) | 489 (75%) | 163 (77%) | 326 (75%) |  |
| Doctor's office (virtual visit) | 9 (1.4%) | 2 (0.9%) | 7 (1.6%) |  |
| Emergency department | 3 (0.5%) | 1 (0.5%) | 2 (0.5%) |  |
| Health clinic or center | 46 (7.1%) | 10 (4.7%) | 36 (8.3%) |  |
| No regular place | 39 (6.0%) | 19 (8.9%) | 20 (4.6%) |  |
| Urgent care | 17 (2.6%) | 6 (2.8%) | 11 (2.5%) |  |
| Other | 8 (1.2%) | 3 (1.4%) | 5 (1.1%) |  |
| I choose not to answer | 37 (5.7%) | 9 (4.2%) | 28 (6.4%) |  |
| Have you received CPR/AED training prior to today? |  |  |  | **<0.001** |
| Yes (Both) | 197 (32%) | 49 (24%) | 148 (36%) |  |
| Yes (CPR Only) | 127 (21%) | 38 (19%) | 89 (22%) |  |
| Yes (AED Only) | 2 (0.3%) | 1 (0.5%) | 1 (0.2%) |  |
| No | 284 (46%) | 117 (57%) | 167 (41%) |  |
| I choose not to answer | 7 (1.1%) | 0 (0%) | 7 (1.7%) |  |
| **Bolded values indicate statistically significant estimates at $p < 0.05$.** |  |  |  |  |

**Table 2.** Self-Reported Barriers of Survey Respondents to Starting CPR & Calling 9-1-1.

|  |  |  |  |  |
| --- | --- | --- | --- | --- |
| Characteristic | Overall (N = 651) | Muslim (N = 214) | Non-Muslim (N = 437) | p-value |
| *Barriers to Starting CPR* |  |  |  |  |
| No problem starting CPR |  |  |  | 0.2 |
| Strongly Disagree | 69 (13%) | 24 (13%) | 45 (13%) |  |
| Disagree | 46 (9.0%) | 16 (9.0%) | 30 (9.0%) |  |
| Neutral | 117 (23%) | 51 (29%) | 66 (20%) |  |
| Agree | 99 (19%) | 34 (19%) | 65 (19%) |  |
| Strongly Agree | 182 (35%) | 53 (30%) | 129 (39%) |  |
| Language Barrier |  |  |  | 0.021 |
| Strongly Disagree | 214 (43%) | 70 (39%) | 144 (44%) |  |
| Disagree | 114 (23%) | 42 (24%) | 72 (22%) |  |
| Neutral | 101 (20%) | 47 (26%) | 54 (17%) |  |
| Agree | 50 (10.0%) | 16 (9.0%) | 34 (10%) |  |
| Strongly Agree | 23 (4.6%) | 3 (1.7%) | 20 (6.2%) |  |
| Concern for Violence |  |  |  | 0.2 |
| Strongly Disagree | 195 (39%) | 59 (33%) | 136 (43%) |  |
| Disagree | 93 (19%) | 39 (22%) | 54 (17%) |  |
| Neutral | 123 (25%) | 51 (29%) | 72 (23%) |  |
| Agree | 51 (10%) | 19 (11%) | 32 (10%) |  |
| Strongly Agree | 34 (6.9%) | 10 (5.6%) | 24 (7.5%) |  |
| Fear of Doing Something Wrong |  |  |  | 0.2 |
| Strongly Disagree | 132 (26%) | 43 (24%) | 89 (27%) |  |
| Disagree | 67 (13%) | 18 (10%) | 49 (15%) |  |
| Neutral | 108 (21%) | 48 (27%) | 60 (18%) |  |
| Agree | 124 (25%) | 42 (23%) | 82 (25%) |  |
| Strongly Agree | 75 (15%) | 28 (16%) | 47 (14%) |  |
| Cost |  |  |  | 0.022 |
| Strongly Disagree | 240 (48%) | 75 (41%) | 165 (53%) |  |
| Disagree | 107 (22%) | 44 (24%) | 63 (20%) |  |
| Neutral | 96 (19%) | 47 (26%) | 49 (16%) |  |
| Agree | 32 (6.5%) | 11 (6.0%) | 21 (6.7%) |  |
| Strongly Agree | 20 (4.0%) | 5 (2.7%) | 15 (4.8%) |  |
| Immigration Status |  |  |  | 0.2 |
| Strongly Disagree | 255 (51%) | 83 (47%) | 172 (53%) |  |
| Disagree | 116 (23%) | 45 (25%) | 71 (22%) |  |
| Neutral | 82 (16%) | 37 (21%) | 45 (14%) |  |
| Agree | 28 (5.6%) | 9 (5.1%) | 19 (5.9%) |  |
| Strongly Agree | 19 (3.8%) | 4 (2.2%) | 15 (4.7%) |  |
| Law Enforcement |  |  |  | <0.001 |
| Strongly Disagree | 207 (41%) | 58 (33%) | 149 (46%) |  |
| Disagree | 121 (24%) | 43 (24%) | 78 (24%) |  |
| Neutral | 102 (20%) | 51 (29%) | 51 (16%) |  |
| Agree | 47 (9.3%) | 20 (11%) | 27 (8.3%) |  |
| Strongly Agree | 27 (5.4%) | 5 (2.8%) | 22 (6.7%) |  |
| *Barriers to Calling 9-1-1* |  |  |  |  |
| No problem calling 9-1-1 |  |  |  | **<0.001** |
| Strongly Disagree | 66 (12%) | 21 (11%) | 45 (12%) |  |
| Disagree | 37 (6.6%) | 11 (5.7%) | 26 (7.1%) |  |
| Neutral | 84 (15%) | 48 (25%) | 36 (9.9%) |  |
| Agree | 98 (18%) | 33 (17%) | 65 (18%) |  |
| Strongly Agree | 274 (49%) | 81 (42%) | 193 (53%) |  |
| Language Barrier |  |  |  | **0.044** |
| Strongly Disagree | 256 (49%) | 83 (43%) | 173 (52%) |  |
| Disagree | 102 (19%) | 41 (21%) | 61 (18%) |  |
| Neutral | 106 (20%) | 51 (26%) | 55 (17%) |  |
| Agree | 39 (7.4%) | 13 (6.7%) | 26 (7.8%) |  |
| Strongly Agree | 23 (4.4%) | 6 (3.1%) | 17 (5.1%) |  |
| Concern for Violence |  |  |  | 0.2 |
| Strongly Disagree | 205 (39%) | 62 (33%) | 143 (43%) |  |
| Disagree | 111 (21%) | 46 (24%) | 65 (20%) |  |
| Neutral | 93 (18%) | 40 (21%) | 53 (16%) |  |
| Agree | 70 (13%) | 25 (13%) | 45 (14%) |  |
| Strongly Agree | 41 (7.9%) | 16 (8.5%) | 25 (7.6%) |  |
| Fear of Doing Something Wrong |  |  |  | **<0.001** |
| Strongly Disagree | 198 (38%) | 59 (31%) | 139 (41%) |  |
| Disagree | 112 (21%) | 42 (22%) | 70 (21%) |  |
| Neutral | 120 (23%) | 63 (33%) | 57 (17%) |  |
| Agree | 66 (13%) | 19 (9.9%) | 47 (14%) |  |
| Strongly Agree | 31 (5.9%) | 8 (4.2%) | 23 (6.8%) |  |
| Cost |  |  |  | **<0.001** |
| Strongly Disagree | 201 (39%) | 52 (28%) | 149 (45%) |  |
| Disagree | 90 (17%) | 29 (15%) | 61 (19%) |  |
| Neutral | 101 (19%) | 55 (29%) | 46 (14%) |  |
| Agree | 70 (14%) | 31 (16%) | 39 (12%) |  |
| Strongly Agree | 56 (11%) | 22 (12%) | 34 (10%) |  |
| Immigration Status |  |  |  | **0.038** |
| Strongly Disagree | 265 (52%) | 82 (44%) | 183 (57%) |  |
| Disagree | 98 (19%) | 41 (22%) | 57 (18%) |  |
| Neutral | 88 (17%) | 42 (22%) | 46 (14%) |  |
| Agree | 30 (5.9%) | 11 (5.9%) | 19 (5.9%) |  |
| Strongly Agree | 27 (5.3%) | 12 (6.4%) | 15 (4.7%) |  |
| Law Enforcement |  |  |  | **<0.001** |
| Strongly Disagree | 222 (41%) | 58 (30%) | 164 (48%) |  |
| Disagree | 106 (20%) | 43 (22%) | 63 (18%) |  |
| Neutral | 95 (18%) | 54 (28%) | 41 (12%) |  |
| Agree | 73 (14%) | 28 (15%) | 45 (13%) |  |
| Strongly Agree | 41 (7.6%) | 10 (5.2%) | 31 (9.0%) |  |
| **Bolded values indicate statistically significant estimates at p < 0.05.** |  |  |  |  |

#### Regressions

##### Barriers to Calling 9-1-1

Religion and race were both significantly associated with law enforcement as a barrier to calling 9-1-1. Compared to Muslim respondents, non-Muslim respondents were 47% less likely to indicate law enforcement as a barrier to calling 9-1-1 (OR: 0.53; 95% CI: 0.29, 0.97; p = 0.04). Compared to respondents who were White or Caucasian, respondents who selected “Other” or “I choose not to answer” were, respectively, 3.24 times (95% CI: 1.18, 9.68; p = 0.03) and 14.72 times (95% CI: 1.85, 506.38; p = 0.04) more likely to indicate law enforcement as a barrier to calling 9-1-1.

Race was significantly associated with immigration status, concern for violence, and fear of doing something wrong. Compared to respondents who were White/Caucasian, the odds of immigration status as a barrier to calling 9-1-1 were 1.98 times higher in Black/African American respondents (95% CI: 1.01, 3.94; p = 0.05) and 2.04 times higher in Hispanic/Latino respondents (95% CI: 0.06, 0.78; p = 0.02). Concern for violence as a barrier to calling 9-1-1 was 2.12 times higher in Black/African American respondents, 2.16 times higher in Hispanic/Latino respondents, 13.68 times higher in multi-/biracial respondents, and 4.07 times higher in respondents who selected “Other”, compared to White/Caucasian respondents.

##### Barriers to Starting CPR

Religion was significantly associated with no problem starting CPR (Table 4). Non-Muslim respondents were 9% less likely to indicate “no problem” compared to Muslim respondents (OR: 0.91; 95% CI: 0.04, 1.80; p = 0.04).

**Table 3.**
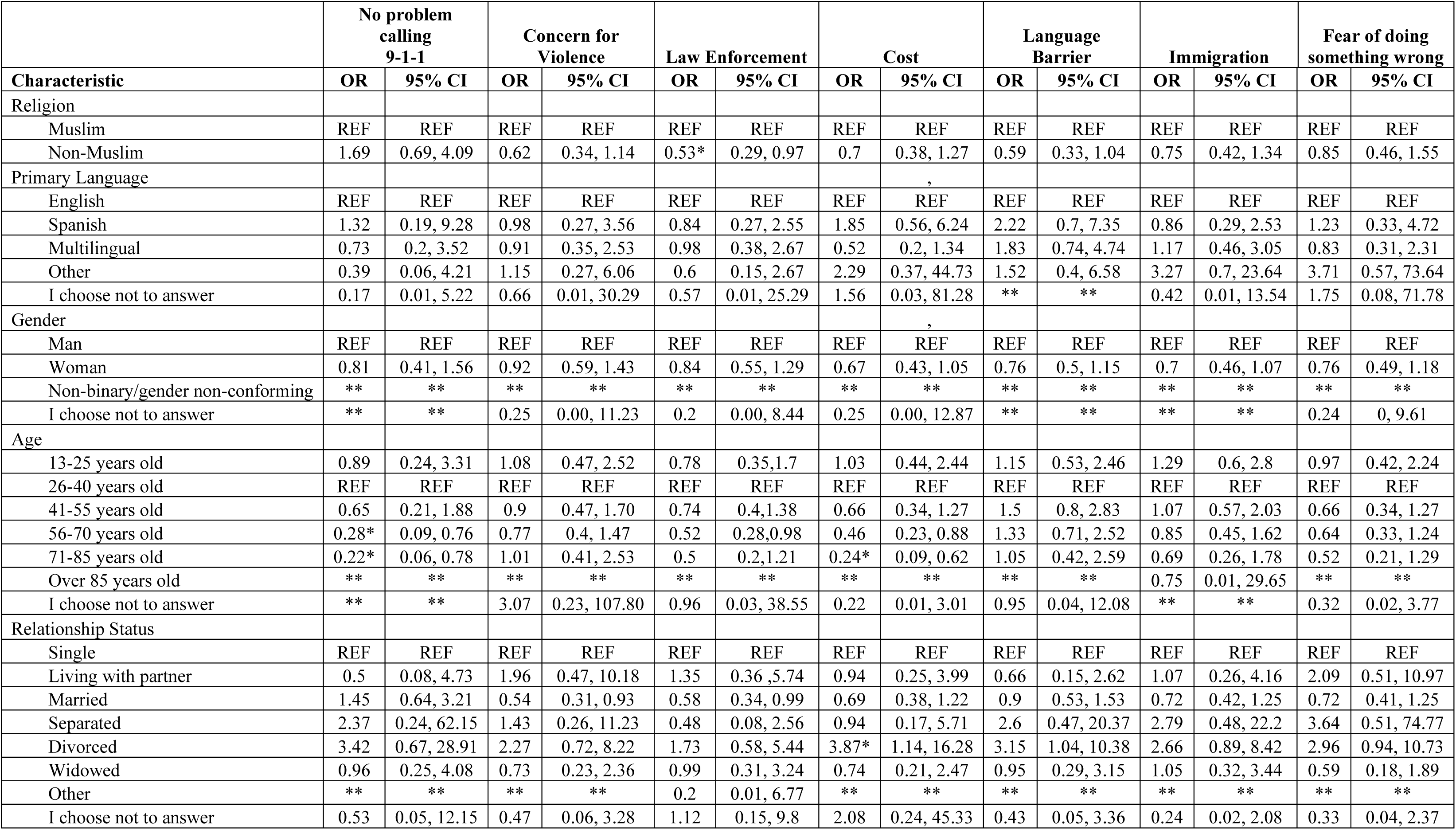
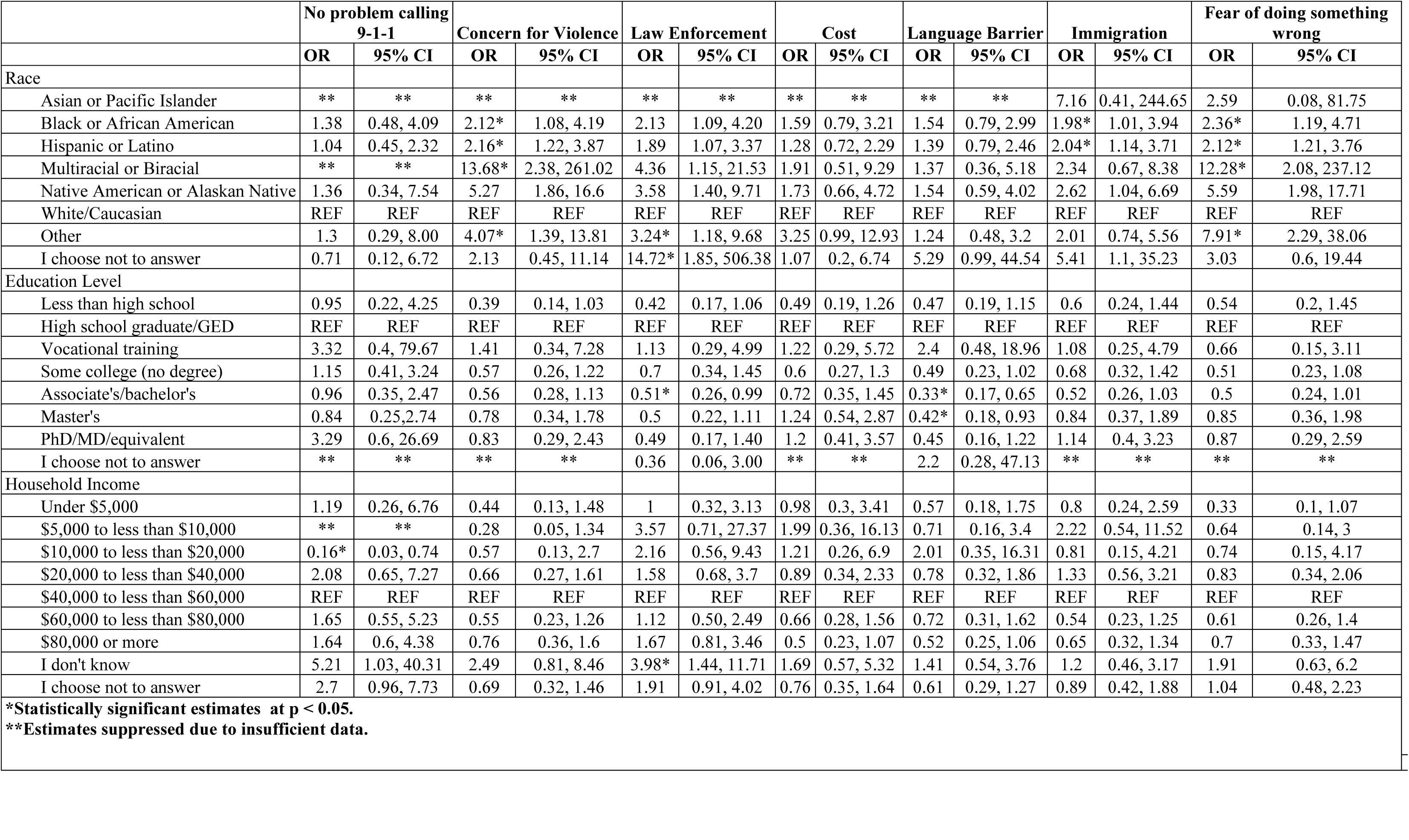
Regression Analysis Between Religion and Barriers to Calling 9-1-1 Among Survey Participants.

**Table 4.**
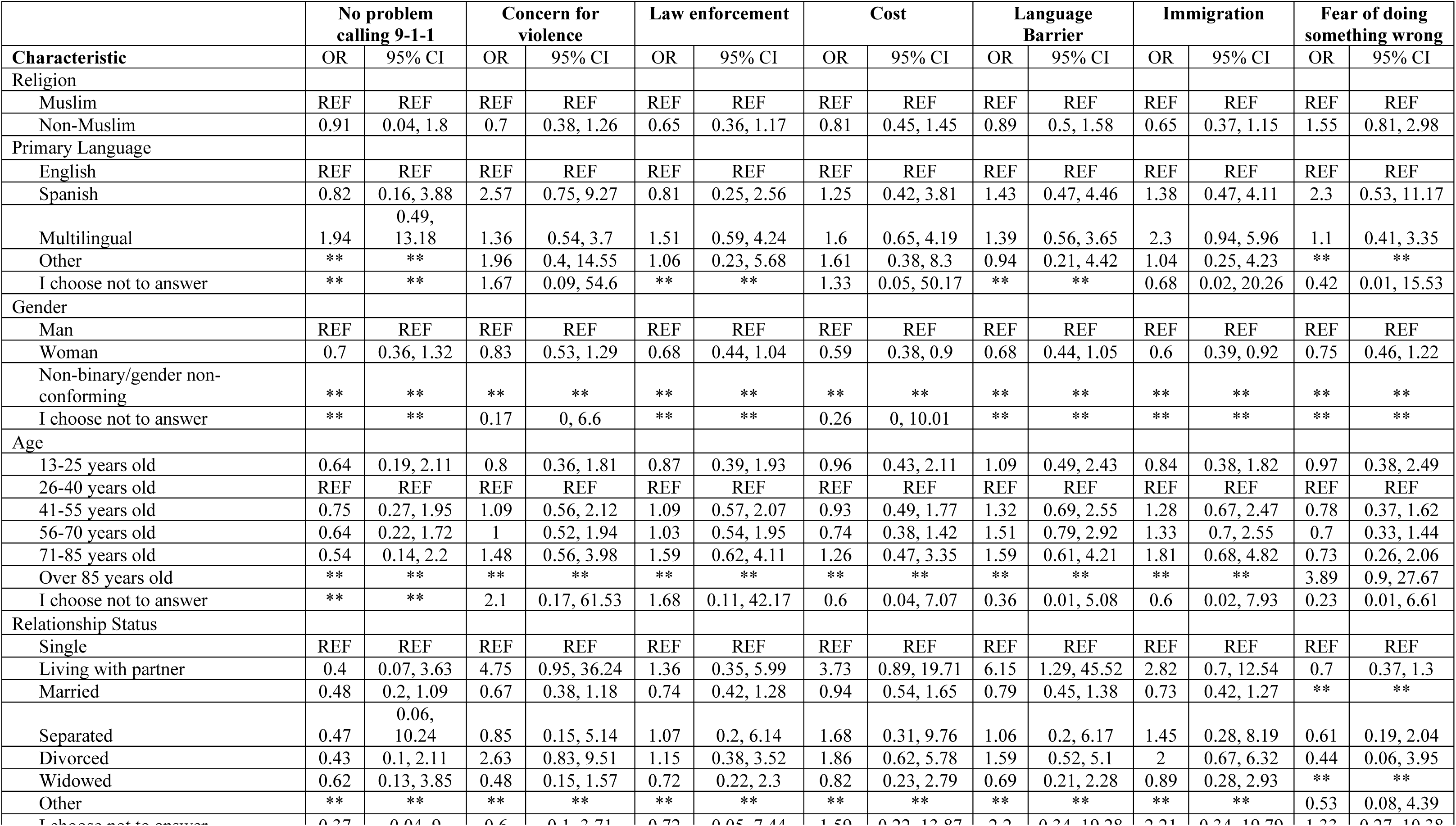
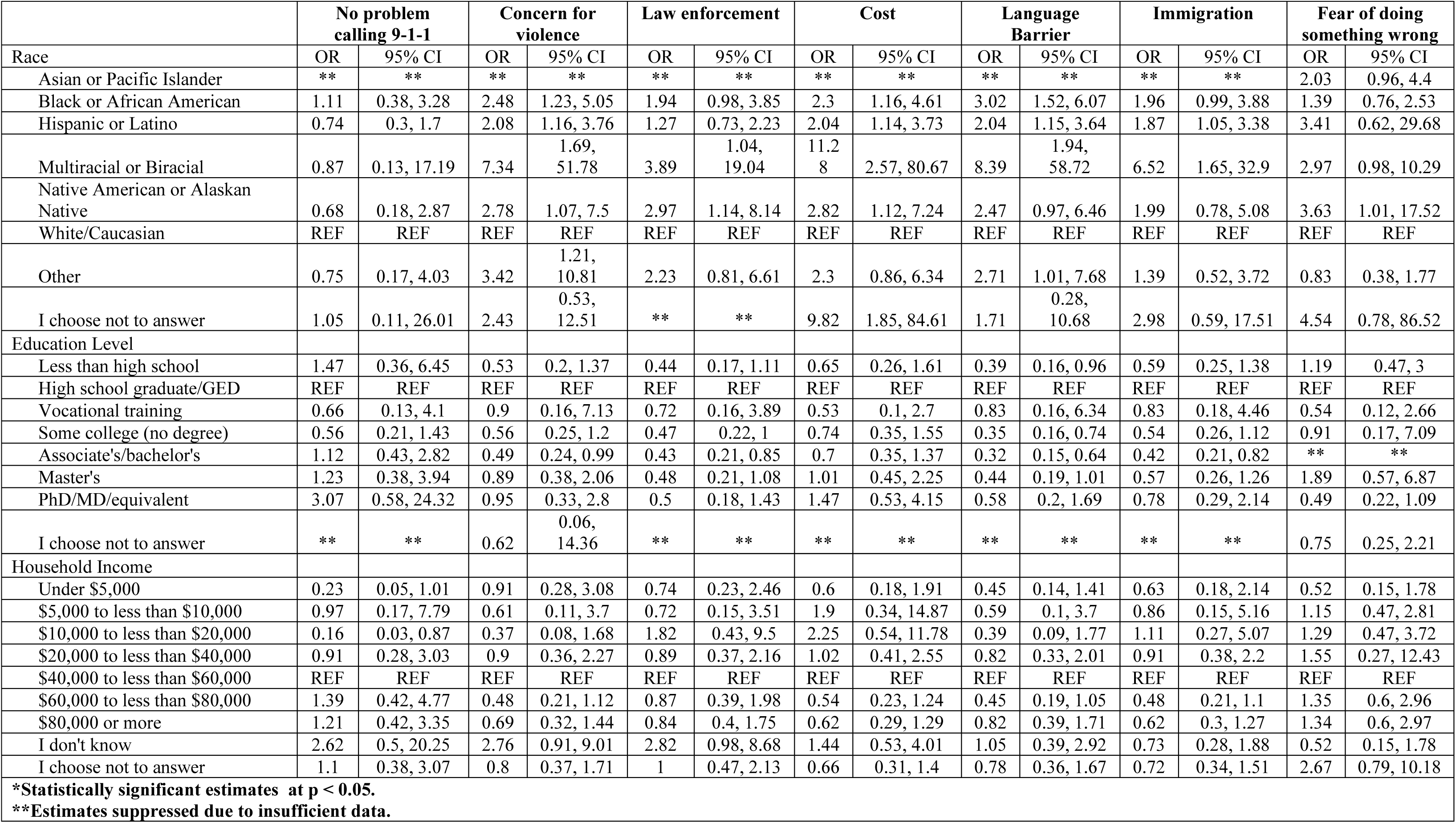
Regression Analysis Between Religion and Barriers to Starting CPR Among Survey Participants.

Race was significantly associated with law enforcement and language as a barrier to starting CPR. Compared to White/Caucasian respondents, Native American/Alaskan Native respondents were 2.97 times more likely to indicate law enforcement as a barrier (95% CI: 1.14, 8.14; p = 0.03). Also compared to White respondents, language as a barrier to starting CPR was 3.02 times higher in Black/African American respondents (95% CI: 1.52, 6.07; p < 0.001), 2.04 times higher in Hispanic/Latino respondents (95% CI: 1.15, 3.64; p = 0.02), and 8.39 times higher in multi-/biracial respondents (95% CI: 1.94, 58.72; p = 0.01).

Gender and race were both significantly associated with immigration status as a barrier to starting CPR. Compared to men, women were 40% less likely (OR: 0.60; 95% CI: 0.39, 0.92; p = 0.02) to indicate immigration status as a barrier to starting CPR. Black/African American (OR: 1.96; 95% CI: 0.99, 3.88; p = 0.05), Hispanic/Latino (OR: 1.87; 95% CI: 1.05, 3.38; p = 0.03), and multi-/biracial (OR: 6.52; 95% CI: 1.65, 32.90; p = 0.01) respondents all had increased odds of indicating immigration status as a barrier to starting CPR, compared to White/Caucasian respondents.

Race was significantly associated with cost and concern for violence as barriers to starting CPR. Compared to White/Caucasian respondents, odds of cost being a barrier to starting CPR increased for respondents who were Black/African American (OR: 2.30; 95% CI: 1.16, 4.61 ; p = 0.02), Hispanic/Latino (OR: 2.04 ; 95% CI: 1.14, 2.73; p = 0.02), Multi-/biracial (OR: 11.28; 95% CI: 2.57, 80.87; p < 0.01), Native American/Alaskan Native (OR: 2.82; 95% CI: 1.12. 7.24; p = 0.03), and respondents who selected “I choose not to answer” (OR: 9.82; 95% CI: 1.85, 84.61; p = 0.01). Also compared to White/Caucasian respondents, odds of concern for violence being a barrier to starting CPR increased for respondents who were Black/African American (OR: 2.48; 95% CI: 1.23, 5.05; p = 0.01), Hispanic/Latino (OR: 2.08; 95% CI: 1.16, 3.76; p = 0.01), Multi-/biracial (OR: 7.34; 95% CI: 1.69, 51.78; p = 0.02), Native American/Alaskan Native (OR: 2.78; 95% CI: 1.21, 10.81; p = 0.04), and respondents who selected “Other” (OR: 3.42; 95% CI: 1.21, 10.81; p = 0.03).

## DISCUSSION

In this study, we noted an important juxtaposition within the Muslim community. Muslim respondents were among the most likely to initiate CPR, demonstrating a strong willingness to act in life-saving situations. Despite this readiness, contacting 9-1-1, the first link in the chain of survival, was a significant barrier for this community. This contrast may reflect the intersection of religious, cultural, and socioeconomic identities within Muslim communities. Factors such as language barriers, perceptions of discrimination, and mistrust of emergency systems may compound to influence how Muslims engage with different aspects of emergency response.

The 2025 AHA Guidelines for Cardiopulmonary Resuscitation and Emergency Cardiovascular Care made specific reference to ‘Disparities and Differences’.^5^ An excerpt from the section states that, “the causes of these disparities are complex and are closely associated with socioeconomic disadvantage and unequal access to care.” This statement broadly characterizes disparities and is a consequence of the paucity of literature exploring unique learner populations, their barriers to CPR education and becoming lay rescuers, and how tailored CPR education should be developed and delivered to effectively empower each distinct learner community to activate the chain of survival. Our previous research explored barriers to CPR education and becoming a lay rescuer in a Spanish community in Southwest Virginia and we found that those learners also were reluctant to contact 9-1-1 and thus initiate the first link in the chain of survival.^20^

Existing literature provides valuable insight into why some Muslims may experience hesitancy in early activation of EMS through calling 9-1-1. Muslim communities often report holding favorable perceptions of their local police; however, attitudes toward law enforcement at the broader societal level tend to be more negative.^29^ These views may be based on factors such as media portrayal and personal experiences with police officers from native countries that have preconceived views.^29^ Other key predictors of negative attitudes toward law enforcement have been identified as fear of discrimination, perceived ethnic bias, and unfavorable media portrayals of Muslims.^22^ These dynamics are further reinforced by documented bias toward Muslims within law enforcement, which intensified in the post 9/11 era and contribute to ongoing mistrust.^30^ Non-White individuals in particular have expressed higher concerns of law enforcement as a barrier to both calling 9-1-1 and starting CPR. In addition, non-White individuals expressed higher concerns of immigration status and concern for violence as barriers. This is consistent with previous research on barriers to calling 9-1-1 or initiating CPR, as well as broader research on attitudes of minority communities.^16–19,31–34^ These reported concerns and barriers among broader minority populations are mirrored in Muslim populations. SDOH such as socioeconomic status, education level, and neighborhood environment significantly shape how Muslims in the U.S. perceive institutional trust.^22^

Collectively, these findings may suggest that delays in early EMS activation stem from broader sociocultural and historical factors, rather than individual-level hesitancy, and may be amplified by structural mistrust and ongoing bias within law enforcement institutions. Current literature highlights opportunities for change through structured diversity initiatives and culturally informed training programs that promote an accurate understanding of Muslim communities and help rebuild trust.^30^

CPR/AED education grounded in religious texts give context and allow learners to view resuscitation efforts in both the lens of medical intervention and a means to fulfill religious obligations. Within Islam, there is an emphasis on lifesaving, reinforced through teachings of the Prophet Muhammad.^35^ Surah 5 (Al-Ma-idah), Verse 2 of the Qur’an states, “And whoever saves a life, it will be as if they saved all of humanity.”^36^ However, Islamic teachings also emphasize modesty and maintain a framework for opposite-sex interactions.^37–38^ Specifically, Muslims believe that opposite gender based interactions should remain limited, unless absolutely necessary.^37^ In order to eliminate barriers and confusion, educational models should address these teachings. Uncertainty regarding the permissibility of initiating CPR or using an AED on individuals of the opposite sex, particularly female victims are all concerns that may arise in the Muslim community.^39^ Addressing religious concerns directly is critical in empowering learners to act in a way that aligns with their religious beliefs.^40^

Addressing barriers to CPR and AED education requires recognition of the socioeconomic disadvantages, racialized identities, and systematic biases faced by Muslim communities, whose identity extends beyond religion alone. Future CPR/AED training for Muslim learners should be tailored to incorporate these diverse identities in a culturally sensitive and inclusive manner, rather than relying on a one-size-fits-all model. As a result of our study, an instructor guide (Appendix B) outlining Islamic perspectives on lifesaving relevancy, opposite-sex interactions, and modesty was created. This document was created and informed through consultation with imams to ensure religious and cultural sensitivity. Creating an environment that empowers Muslim participants to feel safe and confident when acting in the event of a cardiac arrest is critical. Additionally, educational models should emphasize fostering comfort and trust between Muslim communities and law enforcement.

While technical CPR skills and willingness may already be present in this population, CPR/AED educational models must specifically address barriers related to the first link of the chain of survival. Tailored, culturally sensitive education should extend beyond technical instruction to address the needs of Muslim learners, representing a crucial step toward reducing disparities in OHCA outcomes and promoting equity in lifesaving education.

### Limitations

This study should be viewed in light of several limitations. First, interpretability may be limited due to race and ethnicity measurements that did not follow the federal standards by combining race and ethnicity into a single item. Efforts were made to reduce sampling bias by recruiting subjects from similar environments, but it is possible both groups are not fully represented by the sample data. Sample size was determined by feasibility and available participants.

Second, regression models were built to account for some sociodemographic factors, but not all. Sociodemographic factors that were not controlled for could be potential confounders. Prior to the study start, an a priori power analysis was not run. Sample size was determined by feasibility and available participants. As a result, the study may be underpowered to detect small-to-moderate differences across language groups and subgroups, increasing the risk of Type II error and contributing to wide confidence intervals. Despite survey anonymity, the possibility of social desirability bias impacting results with self-reported data is a possible factor to consider.

Third, this study may not account for all barriers faced by Muslim learners. The absence of qualitative data limited the ability to explore context-specific experiences and perceptions that may influence barriers to calling 9-1-1 and initiating CPR within the Muslim population.

Finally, this study did not differentiate between the various branches of Islam, such as Sunni, Shia, Sufi, and smaller sects. Instead, Muslims were considered collectively, which may overlook important nuances in interpretation and practice across these traditions.

## CONCLUSION

Successful activation of the chain of survival and understanding of all the links is key to OHCA survivability. Any community that faces barriers to one or more links of the chain is less prepared to respond to OHCA. Muslim learners were significantly less likely to feel comfortable calling 9-1-1 compared to non-Muslim learners. However, Muslim learners were more likely to have no problem starting CPR. Additional studies are needed to investigate the juxtaposition of comfort levels with these two actions and why there appears to be a disconnect. This study demonstrates the need to further research community-specific barriers in faith communities, specifically faith groups who may be marginalized. In addition, tailored education for the Islamic community must include emphasis on the chain of survival, particularly the importance of calling 9-1-1. Without tailored education, a crucial link is missed in the chain of survival which may result in disruption of the other links in the chain. Further studies are encouraged to understand how religious beliefs, cultural norms, and lived experiences of minority religious and ethnic groups may contribute to barriers to bystander intervention.

## Data Availability

The data that support the findings of this study are available on request from the corresponding author upon reasonable request.

## ACKNOWLEDGEMENTS

The authors wish to thank the Imams who supported this research and assisted with the creation supplemental materials for culturally competent CPR training in Muslim communities.

## References

1. Nolan J, Soar J, Eikeland H. The chain of survival. Resuscitation. 2006;71(3):270–271.

2. Dainty KN, Colquitt B, Bhanji F, Hunt EA, Jefkins T, Leary M, Ornato JP, Swor RA, Panchal A, on behalf of the Science Subcommittee of the American Heart Association Emergency Cardiovascular Care Committee. Understanding the Importance of the Lay Responder Experience in Out-of-Hospital Cardiac Arrest: A Scientific Statement From the American Heart Association. Circulation. 2022;145(17):e852–e867.

3. Marco CA, Larkin GL. Cardiopulmonary resuscitation: Knowledge and opinions among the U.S. general public: State of the science-fiction. Resuscitation. 2008;79(3):490–498.

4. Khanji MY, Waqar S, Khawaja Z, Ali B. How delivering cardiopulmonary resuscitation and basic life support skills training through places of worship can help save lives and address health inequalities. European Heart Journal. 2022;43(24):2257–2260.

5. Del Rios M, Bartos JA, Panchal AR, Atkins DL, Cabañas JG, Cao D, Dainty KN, Dezfulian C, Donoghue AJ, Drennan IR, Elmer J, Hirsch KG, Idris AH, Joyner BL, Kamath-Rayne BD, et al. Part 1: Executive Summary: 2025 American Heart Association Guidelines for Cardiopulmonary Resuscitation and Emergency Cardiovascular Care. Circulation. 2025;152(16_suppl_2):S284–S312.

6. Garcia RA, Spertus JA, Girotra S, Nallamothu BK, Kennedy KF, McNally BF, Breathett K, Rios MD, Sasson C, Chan PS. Racial and Ethnic Differences in Bystander CPR for Witnessed Cardiac Arrest. New England Journal of Medicine. 2022;387(17):1569–1578.

7. Chu K, Swor R, Jackson R, Domeier R, Sadler E, Basse E, Zaleznak H, Gitlin J. Race and Survival After Out-of-Hospital Cardiac Arrest in a Suburban Community. Annals of Emergency Medicine. 1998;31(4):478–482.

8. Brookoff D, Kellermann AL, Hackman BB, Somes G, Dobyns P. Do blacks get bystander cardiopulmonary resuscitation as often as whites? Annals of Emergency Medicine. 1994;24(6):1147–1150.

9. Chen C, Lo CYZ, Ho MJC, Ng Y, Chan HCY, Wu WHK, Ong MEH, Siddiqui FJ. Global Sex Disparities in Bystander Cardiopulmonary Resuscitation After Out-of-Hospital Cardiac Arrest: A Scoping Review. Journal of the American Heart Association. 2024;13(18):e035794.

10. Bradley SM, Fahrenbruch CE, Meischke H, Allen J, Bloomingdale M, Rea TD. Bystander CPR in out-of-hospital cardiac arrest: The role of limited English proficiency. Resuscitation. 2011;82(6):680–684.

11. Nuño T, Bobrow BJ, Rogge-Miller KA, Panczyk M, Mullins T, Tormala W, Estrada A, Keim SM, Spaite DW. Disparities in Telephone CPR Access and Timing During Out-of-Hospital Cardiac Arrest. Resuscitation. 2017;115:11–16.

12. Perera N, Birnie T, Ngo H, Ball S, Whiteside A, Bray J, Bailey P, Finn J. “I’m sorry, my English not very good”: Tracking differences between Language-Barrier and Non-Language-Barrier emergency ambulance calls for Out-of-Hospital Cardiac Arrest. Resuscitation. 2021;169:105–112.

13. CDC. Social Determinants of Health (SDOH). About CDC. 2024.

14. Tsao CW, Aday AW, Almarzooq ZI, Alonso A, Beaton AZ, Bittencourt MS, Boehme AK, Buxton AE, Carson AP, Commodore-Mensah Y, Elkind MSV, Evenson KR, Eze-Nliam C, Ferguson JF, Generoso G, et al. Heart Disease and Stroke Statistics—2022 Update: A Report From the American Heart Association. Circulation. 2022;145(8):e153–e639.

15. Demirovic J. Cardiopulmonary Resuscitation Programs Revisited: Results of a Community Study Among Older African Americans. The American Journal of Geriatric Cardiology. 2004;13(4):182–187.

16. Anderson ML, Cox M, Al-Khatib SM, Nichol G, Thomas KL, Chan PS, Saha-Chaudhuri P, Fosbol EL, Eigel B, Clendenen B, Peterson ED. Cardiopulmonary Resuscitation Training Rates in the United States. JAMA internal medicine. 2014;174(2):194–201.

17. Sasson C, Haukoos JS, Ben-Youssef L, Ramirez L, Bull S, Eigel B, Magid DJ, Padilla R. Barriers to Calling 911 and Learning and Performing Cardiopulmonary Resuscitation for Residents of Primarily Latino, High-Risk Neighborhoods in Denver, Colorado. Annals of Emergency Medicine. 2015;65(5):545–552.e2.

18. Berkowitz D, Cohen JS, McCollum N, Rojas CR, Chamberlain JM. Delays in treatment and disposition attributable to undertriage of pediatric emergency medicine patients. The American Journal of Emergency Medicine. 2023;74:130–134.

19. Anon. US: Immigrants ‘Afraid to Call 911’ | Human Rights Watch.

20. LeNeave C, Meier B, Liffert H, Perkins JC. Impact of Primary Spoken Language as a Social Determinant of Health on Cardiopulmonary Education and Use: Pilot Study. Western Journal of Emergency Medicine: Integrating Emergency Care with Population Health. 2026;27(1).

21. Souaiaia A. What Is the Difference between “Muslim” and “Islamic”? 2016.

22. Yazdiha H. All the Muslims Fit to Print: Racial Frames as Mechanisms of Muslim Ethnoracial Formation in the *New York Times* from 1992 to 2010. Sociology of Race and Ethnicity. 2020;6(4):501–516.

23. Jamaluddin A, Azalea S, Noviar RA, Suwarto DEP, Nugroho NT. The effect of “Mosque Lifesaver Training” on lay persons’ knowledge and willingness to perform basic life support in Indonesia. International Journal of Human and Health Sciences (IJHHS). 2020;5(2):202.

24. Khanji MY, Waqar S, Khawaja Z, Ali B. How delivering cardiopulmonary resuscitation and basic life support skills training through places of worship can help save lives and address health inequalities. European Heart Journal. 2022;43(24):2257–2260.

25. Harris PA, Taylor R, Thielke R, Payne J, Gonzalez N, Conde JG. Research electronic data capture (REDCap)--a metadata-driven methodology and workflow process for providing translational research informatics support. Journal of Biomedical Informatics. 2009;42(2):377–381.

26. Harris PA, Taylor R, Minor BL, Elliott V, Fernandez M, O’Neal L, McLeod L, Delacqua G, Delacqua F, Kirby J, Duda SN, REDCap Consortium. The REDCap consortium: Building an international community of software platform partners. Journal of Biomedical Informatics. 2019;95:103208.

27. R Core Team. R: A Language and Environment for Statistical Computing. Vienna, Austria: R Foundation for Statistical Computing; 2013.

28. Anon. stats package - RDocumentation.

29. Robles J. Islam Is the New Black: Muslim Perceptions of Law Enforcement. McNair Scholars Research Journal. 2017;13(1).

30. Dubosh E, Poulakis M, Abdelghani N. Islamophobia and Law Enforcement in a Post 9/11 World. Islamophobia Studies Journal. 2015;3:138–157.

31. Weitzer R, Tuch SA. Race, Class, and Perceptions of Discrimination by the Police. Crime & Delinquency. 1999;45(4):494–507.

32. Kisa S, Kisa A. “No Papers, No Treatment”: a scoping review of challenges faced by undocumented immigrants in accessing emergency healthcare. International Journal for Equity in Health. 2024;23(1):184.

33. Stadeli KM, Sonett D, Conrick KM, Moore M, Riesenberg M, Bulger EM, Meischke H, Vavilala MS. Perceptions of Prehospital Care for Patients With Limited English Proficiency Among Emergency Medical Technicians and Paramedics. JAMA network open. 2023;6(1):e2253364.

34. Sasson C, Haukoos JS, Bond C, Rabe M, Colbert SH, King R, Sayre M, Heisler M. Barriers and Facilitators to Learning and Performing Cardiopulmonary Resuscitation in Neighborhoods With Low Bystander Cardiopulmonary Resuscitation Prevalence and High Rates of Cardiac Arrest in Columbus, OH. Circulation: Cardiovascular Quality and Outcomes. 2013;6(5):550–558.

35. Masruri M, Al E. The perspective of al-Sunnah al-Nabawiyyah to handle the outbreak of a Pandemic Disease. Turkish Journal of Computer and Mathematics Education (TURCOMAT). 2021;12(2):686–693.

36. Abdel Haleem MA ed. The Qurʾan. New York: Oxford University Press; 2005.

37. Haris AR, Syayfi S, Nurbaiti T. Thematic Study of the Concept of Interaction between Opposite Sexes in the Qur’an. International Journal of Multidisciplinary Research and Growth Evaluation. 2023;4(3):1044–1048.

38. Jneid M. Gender Interaction in Islam. Muslim World Research Centre.

39. Peña-Pinedo G, Hernández-Patiño I. CPR knowledge in future physicians: evidence to strengthen undergraduate medical training. Frontiers in Medicine. 2025;12.

40. Padela AI, Pozo PR del. Muslim patients and cross-gender interactions in medicine: an Islamic bioethical perspective. Journal of Medical Ethics. 2011;37(1):40–44.

